# Epidemiology of COVID-19 infection amongst workers in Primary Healthcare in Qatar

**DOI:** 10.1101/2021.02.15.21251586

**Authors:** Mohamed Ghaith Al-Kuwari, Mariam AbdelMalik, Asma Ali Al-Nuaimi, Jazeel Abdulmajeed, Hamad Eid Al-Romaihi, Sandy Semaan

**Author notes:** **Correspondence author:** Sandy Semaan, QIPCO Compound, Villa 241 Lusail, Qatar.

## Abstract

**Background:** COVID-19 transmission was significant amongst Healthcare workers worldwide.

**Aim:** This study aims to estimate the risk of exposure for COVID-19 across Primary Healthcare workers in the State of Qatar. Methods: A cross-sectional descriptive study was conducted to study the burden of COVID-19 among staff working at PHCC during the COVID-19 pandemic from March 1 to October 31, 2020.

**Results:** 1,048 (87.4%)of the infected HCWs belonged to the age group below 45 years, and 488 (40.7%) HCWs were females. 450 (37.5%) were HCWs clinical staff working in one of the 27 PHCC HCs; Despite the increased patient footfall and risk environment, the COVID HCs had an attack rate of 10.1%, which is not significantly different from the average attack rate of 8.9% among staff located in other HCs (p-value =0.26). Storekeepers, engineering & maintenance staff, housekeeping staff, support staff, and security staff (outsourced positions) had the highest positivity rates, 100%, 67.2%, 47.1%, 32.4%, and 29.5% respective positivity rates.

**Conclusions:** The elevated risk of infection amongst outsourced healthcare workers can be explained by environmental factors such as living conditions. On the other hand, better containment within clinical healthcare workers can be attributed to strict safety training and compliance with preventative measures which is recommended to be implemented across all settings.

## Introduction

COVID-19 disease has affected more than 100 million individuals worldwide. Health care workers (HCW) are at increased risk of contracting infectious diseases because of their occupational exposure.^1^

In the State of Qatar, more than 150,000 persons were infected with around 200 deaths as of January 2021 ^2^. Qatar has taken many public health measures such as social distancing strategies to protect its population from COVID-19 disease and to reduce the incidence of new cases, mostly that no specific pharmaceutical intervention was available during the first surge of the pandemic in 2020.^3^

As part of the State of Qatar’s efforts to control the COVID-19 pandemic, Primary Healthcare Corporation (PHCC) has had a frontline presence and a proactive role in reducing the spread of coronavirus in Qatar, with dedicated COVID-19 Center, contact tracing, and dedicated Drive Through swabbing hubs to assist with early detection.^4^ The Corporation comprises a network of 27 health centers and employs more than 6,000 employees.

International studies have also estimated that frontline healthcare workers had a higher risk than people living in the general community of reporting a positive test, adjusting for the likelihood of receiving a test,^5,6^ and the prevalence of exposed workers in the healthcare Industry^7,8^.

A National study in Qatar has identified that COVID-19 infection often occurs with HCWs who are not directly working with COVID-19 patients. One of the reasons depicted is that PPE use is less stringent in such settings ^9^

However, there is still limited information available about COVID-19 epidemiological characteristics among HCW, and it varies in different geographical regions of the world.^10,11^ Understanding the epidemiology of COVID-19 infection among healthcare workers at Primary Care Settings is a crucial factor in determining the outbreak trajectories and clinical outcomes at the population level, considering their extent of interaction with the health seeking population in times of a health emergency.

In this study, we aim to estimate the burden of COVID-19 infection amongst all types of workers active at Primary Health Care Corporation and identify specific health care workgroups who may be particularly vulnerable to the disease during the ongoing COVID-19 pandemic.

## Methodology

A cross-sectional descriptive study was conducted to study the burden of COVID-19 among health care workers (HCW) working at PHCC during the COVID-19 pandemic. All HCWs who tested positive for COVID-19 during the period from March 1 to October 31, 2020, were included for analysis

### Definitions

For this study, a Healthcare Worker was defined as any person serving in a Primary Healthcare Corporation healthcare setting, either directly hired or a contractual employee, who had the potential for direct or indirect exposure to patients or their infectious secretions and materials, including, but not limited to, physicians, nurses, paramedics, laboratory workers, clinical support staff, e.g., wellness gym instructor, administrative staff, facility officer, security officer or maintenance workers.

### Data sources

Secondary data available from PHCC databases were compiled and utilized for this study. Data were extracted from the PHCC staff database, including demographics of the personnel, work location during the pandemic, and other related information. Subsequently, this data was mapped to the COVID-19 PCR results available on the electronic medical record (Cerner).

The compiled data extract was imported into STATA v 15.1 – (StataCorp. 2017. College Station, TX: StataCorp LLC.). Chi–square test or Fisher’s exact test was used as appropriate; a p-value of <0.05 was considered significant.

The attack rate (AR) was calculated as the percentage of the cumulative number of laboratory-confirmed COVID-19 positive HCW divided by the total number of HCWs. The test positivity (PR) was defined as the percentage of the cumulative number of laboratory-confirmed COVID-19 positive HCW divided by the total number of HCWs tested.

## Results

During the study period extending from March 1 to October 31, 2020 Primary Healthcare Corporation (PHCC) has employed 9,172 staff. Among the 7,407 (81%) staff who were subjected to COVID-19 RT-PCR tests, 1,199 (16.2%) were positive. An overall attack rate of 13.1% was estimated.

The first case among PHCC staff was detected on March 12^th^, 2020 (week 12). A major peak of cases was observed during April May (week 18-19), as shown in Figure 1.

**Figure 1.**
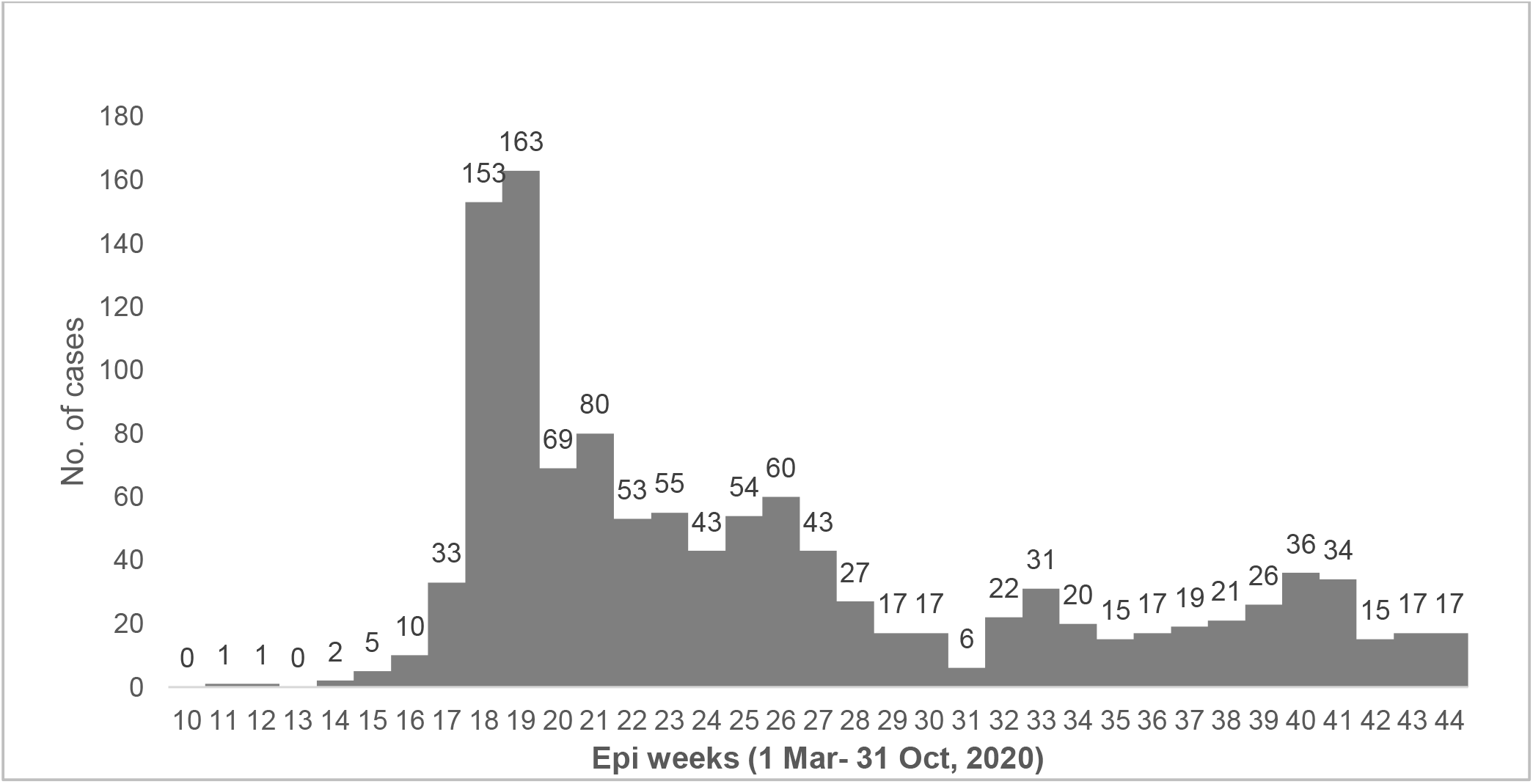
Epidemic curve with case number of HCWs with COVID-19 in PHCC from 1 March to 31 October 2020 (Epiweeks 10-44).

The median age of the infected HCWs was 36 years. 1,048 (87.4%) belonged to the age group below 45 years and 488 (40.7%) HCWs were females. 695 (58%) were directly hired regular employees of PHCC, while 450 (37.5%) HCWs were clinical staff working in one of the 27 PHCC HCs; amongst them 131 (10.9% of the infected HCWs) worked in the 4 designated COVID-19 Health Centre.

Significant difference was observed in the positivity rates while comparing the infected HCWs based on various variables. HCWs aged less than 45 years had a higher attack rate (14.5%) and test positivity (17.5%) compared to their colleagues aged above 45 years (*p value <0*.*001*). Male employees had a higher attack rate (18.5%) and test positivity (23.8%) compared to female employees (*p value <0*.*001*). Non-clinical occupations had a higher attack rate (19.7%) and test positivity (26.8%) compared to clinical occupations (*p value <0*.*001*). Contractual employees had a higher attack rate (42.9%) and test positivity (44.4%) compared to regular PHCC employees (*p value <0*.*001*).

No significant difference was observed in the infection rates of employees who worked in COVID health centers compared to those working in other PHCC health centers (*p value 0*.*61*). Detailed comparison of estimated rates is provided in table 1.

**Table 1:** PHCC Staff Characteristics, screening proportion, attack rate and positivity rate (1 March to 31 October)

| VARIABLE | TOTAL<br>STAFF | TESTED | POSITIVES | ATTACK<br>RATE | TEST<br>POSITIVITY | *p value* |
| --- | --- | --- | --- | --- | --- | --- |
| ALL STAFF | 9,172 | 7,407 | 1,199 | 13.1% | 16.2% |  |
| AGE GROUP |  |  |  |  |  | <0.001 |
| less than 45y | 7,250 | 5,999 | 1,048 (87.4%) | 14.5% | 17.5% |  |
| 45y and above | 1,922 | 1,408 | 151 (12.6%) | 7.9% | 10.7% |  |
| GENDER |  |  |  |  |  | <0.001 |
| Female | 5,320 | 4,415 | 488 (40.7%) | 9.2% | 11.1% |  |
| Male | 3,852 | 2,992 | 711 (59.3%) | 18.5% | 23.8% |  |
| OCCUPATION |  |  |  |  |  | <0.001 |
| Clinical | 5,363 | 4,610 | 450 (37.5%) | 8.4% | 9.8% |  |
| Non Clinical | 3,809 | 2,797 | 749 (62.5%) | 19.7% | 26.8% |  |
| TYPE OF<br>EMPLOYMENT |  |  |  |  |  | <0.001 |

## Clinical vs Non-clinical staff

Among the clinical staff, all occupations have been affected by the spread of the COVID-19 with positivity rates ranging between approximately 6% and 12%. In particular, pharmacists, dentists, wellness gym staff, and nurses have had higher positivity rates compared to the others-12.7%, 11.2%, 10.7% and 10.5% respectively.

Amongst the non-clinical occupations, storekeepers, engineering & maintenance staff, housekeeping staff, support staff, and security staff had the highest positivity rates, 100%, 67.2%, 47.1%, 32.4%, and 29.5% respective positivity rates (Table 3). Administrative Staff who are predominant amongst non-clinical staff had a positivity rate of 3.5% and an attack rate of 1.8%.

**Table 2.**
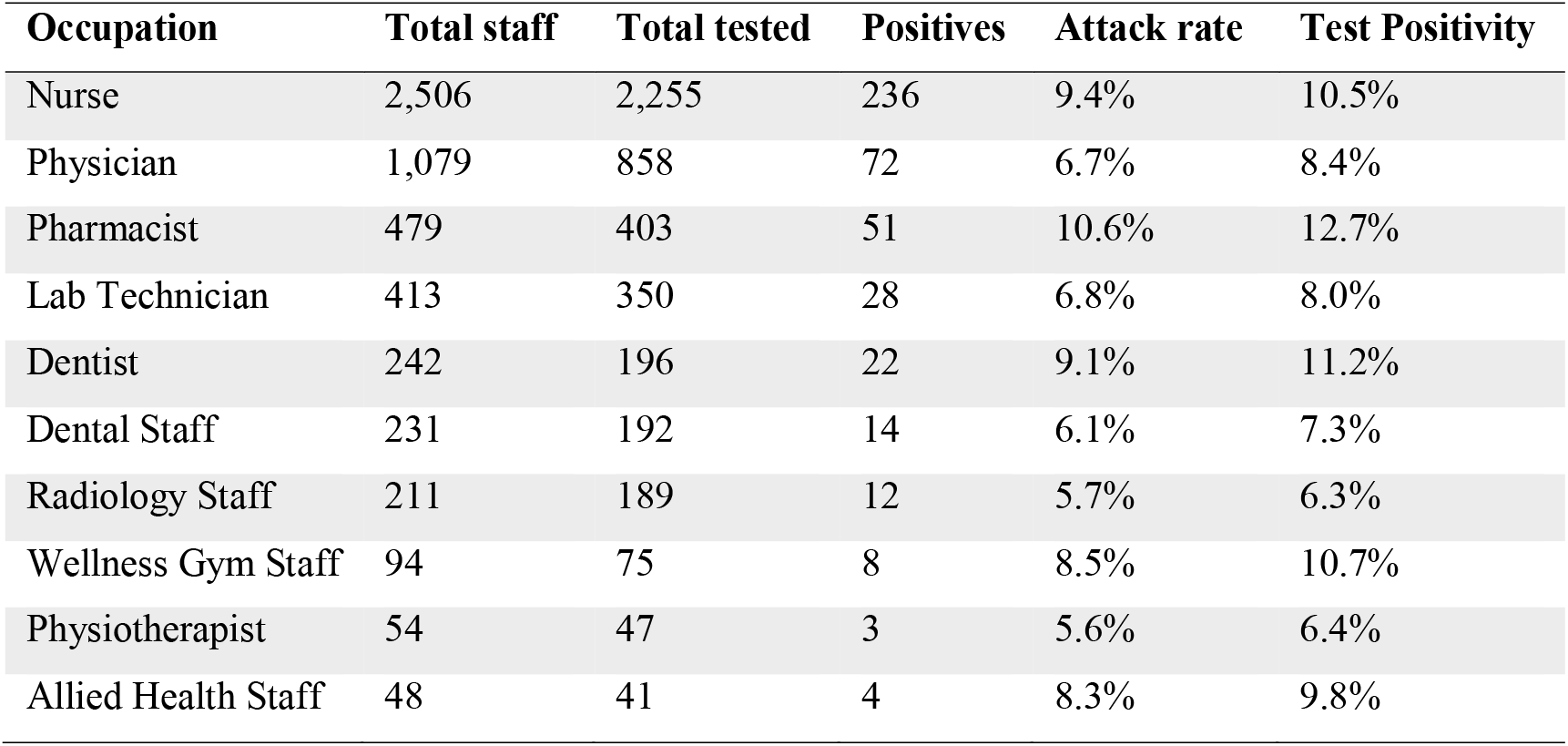
Attack rate and Test positivity among clinical staff

| Occupation | Total staff | Total tested | Positives | Attack rate | Test Positivity |
| --- | --- | --- | --- | --- | --- |
| Nurse | 2,506 | 2,255 | 236 | 9.4% | 10.5% |
| Physician | 1,079 | 858 | 72 | 6.7% | 8.4% |
| Pharmacist | 479 | 403 | 51 | 10.6% | 12.7% |
| Lab Technician | 413 | 350 | 28 | 6.8% | 8.0% |
| Dentist | 242 | 196 | 22 | 9.1% | 11.2% |
| Dental Staff | 231 | 192 | 14 | 6.1% | 7.3% |
| Radiology Staff | 211 | 189 | 12 | 5.7% | 6.3% |
| Wellness Gym Staff | 94 | 75 | 8 | 8.5% | 10.7% |
| Physiotherapist | 54 | 47 | 3 | 5.6% | 6.4% |
| Allied Health Staff | 48 | 41 | 4 | 8.3% | 9.8% |

**Table 3.** Attack rate and Test positivity among non-clinical staff

|  | Total staff | Total tested | Positives | Attack rate | Test Positivity |
| --- | --- | --- | --- | --- | --- |
| Administrative staff | 1326 | 689 | 24 | 1.8% | 3.5% |
| Receptionists and cashiers | 807 | 657 | 112 | 13.9% | 17.0% |
| Housekeeping staff | 530 | 526 | 248 | 46.8% | 47.1% |
| Support staff | 390 | 358 | 116 | 29.7% | 32.4% |
| Security officers | 388 | 322 | 95 | 24.5% | 29.5% |
| Transport staff | 135 | 82 | 18 | 13.3% | 22.0% |
| Customer service staff | 106 | 81 | 10 | 9.4% | 12.3% |
| Engineering & maintenance staff | 112 | 67 | 45 | 40.2% | 67.2% |
| Storekeeper | 15 | 15 | 15 | 100% | 100% |

Out of the 27 PHCC Health Centers (HC), 4 HCs had been designated official assessment and triage COVID-19 Health centers starting March 15, 2020 (Table 4). Similar patient volumes were seen at both these categories of HCs-143,154 suspected patients were swabbed at the 4 COVID HCs and a cumulative number of 145,565 suspected patients were swabbed in all other HCs.

**Table 4.**
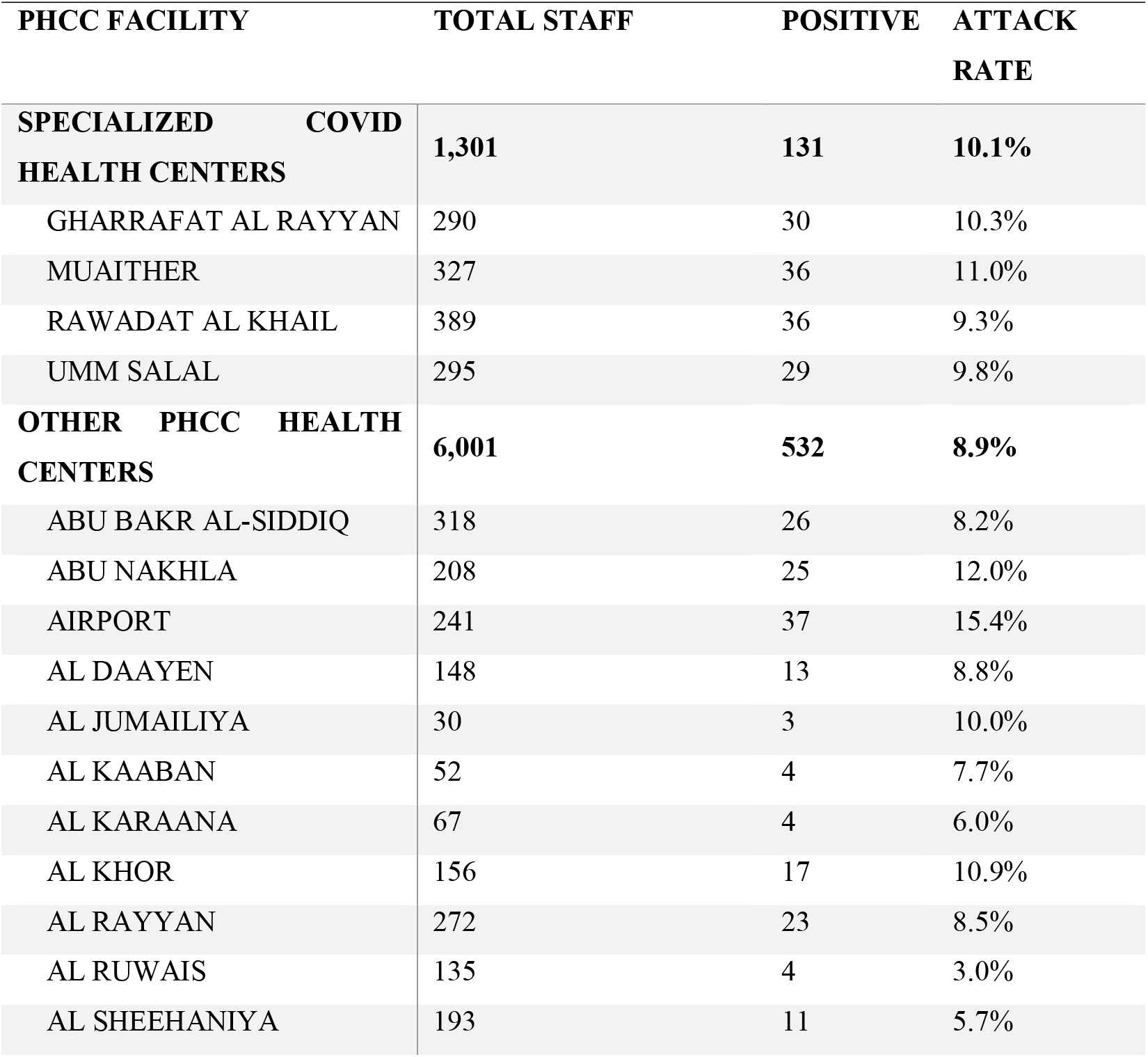
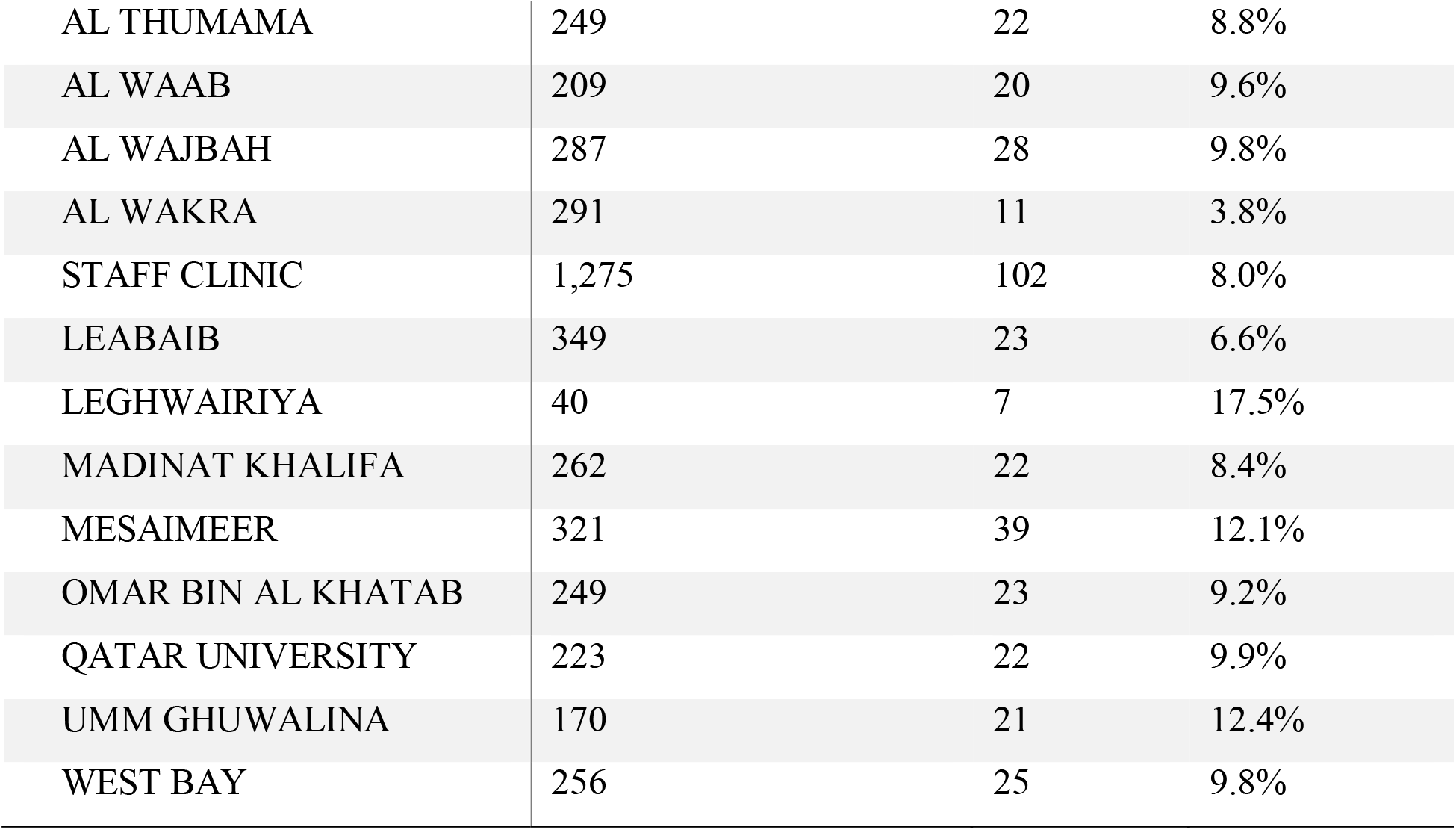
Attack rate among staff in various health centers

Despite the increased patient foot fall and risk environment, the COVID HCs had an attack rate of 10.1%, which is not significantly different from the average attack rate of 8.9% among staff located in other HCs (*p value =0*.*26*)

## Discussion

Primary HealthCare Corporation has taken precautionary measures to prevent the spread of COVID-19 amongst its healthcare workers. PHCC Health Centers have maintained vital services provided to patients such as well-baby and vaccinations, ultrasound, and premarital testing clinics, all by encouraging patients to visit health centers only if medical consultation is imperative. Online health services through virtual consultations were provided by PHCC to minimize the risk of exposure and contamination for both patients and medical staff.^12^

Designated assessment and triage COVID-19 centers, although having swabbed almost six times more suspected patients per HC center than other PHCC Health Centers, have seen almost similar attack rates amongst their staff than other PHCC Health Centers. The similarities in the frequency of infected staff, despite the vast difference in the levels of exposure, can be attributed to continuous training and raising awareness amongst staff on the proper use of PPE and the implementation of stringent infection prevention and control policies and procedures which helps prevent the spread of COVID-19 virus. ^**Error! Bookmark not defined**.,13^

PHCC has provided adequate education and training content, which includes the use of personal protective equipment, hand hygiene, medical waste management, sterilization of patient-care devices, and management of occupational exposure. Within these health centers, non-clinical staff who are predominantly outsourced employees seem to have a higher test positivity and attack rates than clinical staff.

The higher positivity and attack rates amongst non-clinical staff could be due to several educational, social, and environmental factors such as lack of awareness and training on how to use PPEs, less enforcement of occupational safety measures, and crowded accommodations, which is considered to be one of the strong forecasters and substantial contributing risk factors for health problems amongst workers^14^. Indeed, craft and manual workers are more likely to live in crowded shared accommodation in constant proximity to one another, increasing the likelihood of COVID-19 spread through community transmission. They also often gather for social and recreational activities, shared dining, and use of shared equipment. ^15^

The lower positivity among clinical staff can be attributed to the stringent enforcement of infection prevention and control measures, despite the front line aspect of their daily work routine ^16^.

Some of these measures include continuously wearing masks, frequent handwashing, and constant availability of sanitizers, in addition to the implementation of social distancing strategies. The administrative staff is considered outliers to the non-clinical staff with low attack rate because they undergo similar safety training as clinical staff and who are more likely to live in separate accommodation. Among the clinical workers, pharmacists, dentists, nurses, and wellness staff have encountered slightly higher positivity rates, which can be attributed to their nature of work as dedicated COVID-19 swabbing staff. Additionally, Pharmacists have frequent dealings with Storekeepers and Q-Post drivers to distribute medication for home delivery. At the same time, the dental team faces a higher risk of infection due to the oral nature of their work. Nurses and wellness staff have daily close encounters with patients and staff alike, being the 1st line of contact in the triage selection process.

Although female staff at PHCC outnumber their male counterpart, the spread of the COVID-19 virus has been more pronounced amongst males with higher positivity and attack rates. Some occupations such as Storekeeper, Security, and Engineering & Maintenance, is predominantly taken up by male staff, have seen considerably high rates of infection. Furthermore, male craft and manual workers, as previously mentioned, are more likely to contract the COVID-19 virus due to the nature of their accommodation and their socio-recreational activities.

According to the collected data, staff below 45 years of age have seen higher positivity and attack rates. The higher positivity and attack rate amongst this age range is mainly because most of the non-clinical outsourced staff are below 45 years of age, in addition to some of the workers above 55 years of age being allowed to work from home and minimize their daily exposure to the virus through a range of teleconsultation services. services.^17^

In evaluating the transmission of COVID-19 amongst hospital staff, it is crucial to test both clinical and non-clinical staff during the pandemic to frame the extent of viral spread. Even with limited infection control measures in non-clinical areas, COVID-19 virus transmission did not occur among hospital staff beyond community outbreak, reflecting the effectiveness of infection control measures and appropriate usage of personal protective equipment^18,19^. This also highlights the need to implement the same stringent control measures to non-clinical staff as well, namely outsourced workers, who should undergo training on how to avoid the spread of the virus by taking proper precautionary measures and making appropriate use of their protective equipment. Improvements in their living conditions will ultimately reduce the risk of infection by promoting social distancing and minimizing community transmissions.

These findings highlight the importance of developing a clear and concise National Occupational Health Policy underscoring the importance of training and infection control measures and outlining minimum requirements of living conditions of staff (direct and outsourced) working in a healthcare setting.

## Data Availability

The datasets analyzed during the current study are available from the corresponding author on
reasonable request.

## Competing Interests

Non Applicable

## Funding

Non Applicable

## Notes

### Competing Interest Statement

The authors have declared no competing interest.

### Funding Statement

Primary Healthcare Corporation will be our funding agency for our research paper

### Author Declarations

Primary HEalthcare Corporation Research Committee have given an ethical approval to our Research Paper on September 6th, 2020.

